# COVID-19 in non-hospitalised adults caused by either SARS-CoV-2 sub-variants Omicron BA.1, BA.2, BA.5 or Delta associates with similar illness duration, symptom severity and viral kinetics, irrespective of vaccination history

**DOI:** 10.1101/2022.07.07.22277367

**Authors:** Hermaleigh Townsley, Joshua Gahir, Timothy W Russell, Edward J Carr, Matala Dyke, Lorin Adams, Murad Miah, Bobbi Clayton, Callie Smith, Mauro Miranda, Harriet V Mears, Chris Bailey, James RM Black, Ashley S Fowler, Margaret Crawford, Katalin Wilkinson, Matthew Hutchinson, Ruth Harvey, Nicola O’Reilly, Gavin Kelly, Robert Goldstone, Rupert Beale, Padmasayee Papineni, Tumena Corrah, Richard Gilson, Simon Caidan, Jerome Nicod, Steve Gamblin, George Kassiotis, Vincenzo Libri, Bryan Williams, Sonia Gandhi, Adam J Kucharski, Charles Swanton, David LV Bauer, Emma C Wall

## Abstract

**Background:** SARS-CoV-2 variant Omicron rapidly evolved over 2022, causing three waves of infection due to sub-variants BA.1, BA.2 and BA.4/5. We sought to characterise symptoms and viral loads over the course of COVID-19 infection with these sub-variants in otherwise-healthy, vaccinated, non-hospitalised adults, and compared data to infections with the preceding Delta variant of concern (VOC).

**Methods:** In a prospective, observational cohort study, healthy vaccinated UK adults who reported a positive PCR or lateral flow test, self-swabbed on alternate days until day 10. We compared symptoms and viral load trajectories between infections caused by VOCs Delta and Omicron (sub-variants BA.1, BA.2 and BA.4/5), and tested for relationships between vaccine dose, symptoms and PCR Ct value as a proxy for viral load.

**Results:** 555 infection episodes were reported among 483 participants. Across VOCs, symptom burden and duration were similar, however symptom profiles differed among infections caused by Delta compared to Omicron sub-variants; symptoms of all Omicron sub-variants BA.1, BA.2 and BA.4/5 were very similar. Anosmia was reported in 7-13% of participants with Omicron sub-variants, compared to 25/60 (42%) with Delta infection (P= 1.31e-08 or 1.03e-05 or 5.63e-05; χ^2^ test d2+Delta vs. Omicron BA.1 or vs. BA.2, or BA.5, respectively), fever was more common with Omicron BA.5 (30/55, 55%) than Delta (20/60, 33%) (p 0.03). Amongst infections with all Omicron sub-variants, symptoms of coryza, fatigue, cough and myalgia predominated. Viral load trajectories and peaks did not differ between Delta, and Omicron, irrespective of symptom severity (including asymptomatic participants), VOC or vaccination status. Ct values were negatively associated with time since vaccination in participants infected with BA.1; however, this trend was not observed in BA.2/BA.4/5 infections.

**Conclusion:** Our study emphasises both the changing symptom profile of COVID-19 infections in the Omicron era, and ongoing transmission risk of Omicron sub-variants in vaccinated adults.

**Trial registration:** NCT04750356

## Introduction

COVID-19 causes a wide range of symptoms in humans; recognition of this diversity now forms the core of global public health messaging, including in the United States, where the Centers for Disease Control and Prevention (CDC) recognise a set of eleven possible symptoms (CDC, 2022) (1). The emergence of new SARS-CoV-2 variants of concern (VOCs) with substantially different properties such as innate immune antagonism (2) and tissue tropism (3, 4), has occurred despite widespread vaccination that induces durable immune responses to these variants (5-7). While vaccination has led to dramatic reductions in hospitalisation and deaths from COVID-19, infection and transmission are less affected by vaccination (8-12). High numbers of COVID-19 cases caused by the Omicron BA.1 and BA.2 sub-variants in vaccinated individuals were reported across national surveillance systems between December 2021-May 2022 (13), despite a major booster vaccination campaign.

Early reports of Omicron BA.1 infection from South Africa in December 2021 suggested this VOC caused a less-severe clinical disease, as measured by crude outcomes of hospitalisation and mortality rates; in the context of highly vaccinated European populations, similar trends have been reported (8, 9, 14). While these data are encouraging, they do not account for the significant ongoing-impact of community COVID-19 infections in non-hospitalised adults, with attendant risks of onward transmission and burden on healthcare, particularly for clinically extremely vulnerable individuals (CEV)(15) and those developing post-COVID syndrome (PCS) (16-18). One study of household transmission in the UK found that while viral kinetics were altered by vaccination, secondary attack rates were similar across VOCs(11). Furthermore, few studies have prospectively examined relationships between symptoms and VOC infection in non-hospitalised adults, reporting changes in symptoms of COVID-19 between VOCs, but have neither captured asymptomatic infections or nor controlled for time since last vaccine dose, and thus waning immunity(11, 19, 20).

The UK’s NHS COVID-19 guidance was changed in April 2022, following the CDC in expanding the list of cardinal symptoms, advising self-isolation based on the presence of fever or illness severity, and defined shorter recommended isolation periods (5 days, with advice to avoid large crowds or contact with clinically-vulnerable individuals for up to 10 days)(1, 21). The guidance also removed the explicit/general recommendation for the use of non-pharmaceutical interventions (NPIs) and testing following COVID symptoms in parallel with withdrawal of free tests. Reduced access to free testing places an increased burden of responsibility on individuals to self-diagnose SARS-CoV-2 infection and take action to prevent infection of contacts. To investigate if symptom-based guidance remains appropriate for Omicron BA.1, BA.2 and BA.5, we compared symptom profiles and viral load trajectories between healthy, vaccinated adults infected with SARS-CoV-2 variants Delta, BA.1, BA.2 and BA.5 stratifying the cohorts by vaccine doses and time since last dose.

## Materials and Methods

### Clinical cohort

We analysed data from participants in the University College London Hospitals (UCLH)-Francis Crick Institute *Legacy* study cohort (NCT04750356), who reported a positive SARS-CoV-2 test either through asymptomatic screening or symptom-based testing. The *Legacy* study was established in January 2021 to track serological responses to vaccination during the national COVID-19 vaccination programme in a prospective cohort of healthy staff volunteers. All participants at the time of recruitment were undergoing mandatory weekly or twice weekly occupational health testing for COVID-19(22). Infection episodes were defined as a positive SARS-CoV-2 test, either through asymptomatic occupational screening or following additional symptomatic testing. Participants reporting an infection episode had same-day swabs collected by courier on alternate days up to day 10 post symptom onset (defined as first day of any symptoms of any severity) or day 10 post positive swab, whichever was earlier. An additional swab where possible was performed between day 11 and day 30 after return to work (Figure 1). Symptom severity in non-hospitalised adults was self-reported via an online symptom diary during the infective period. Participants recorded individual symptom detail, including severity and duration that were cross-checked with a study clinician at a post-infection study visit. To capture the scale of symptom severity experienced by participants, we assigned symptom severity categories to those with asymptomatic infection (0), mild (I), moderate (II) and severe (III), expanding the WHO categories 1-2(23), in the absence of validated severity scores for non-hospitalised adults. Symptoms were defined as follows, grade I: “does not interfere with the participant’s daily routine and does not require further procedure; it causes slight discomfort”; grade II: “interferes with some aspects of the participant’s routine, or requires further procedure, but is not damaging to health; it causes moderate discomfort”; grade III: “results in alteration, discomfort or disability which is clearly damaging to health”. We further categorised symptom profiles in two ways, excluding individuals who had not completed a symptom diary. Firstly, into three categories based on the original NHS symptoms of COVID-19: symptomatic with one or more “classic” cardinal NHS symptoms (cough, fever, anosmia), symptomatic with only non-cardinal symptoms, or not symptomatic. Secondly, into four categories using the updated NHS & CDC guidance(1, 24) on the triggers for isolation with symptomatic SARS-CoV-2: asymptomatic, symptomatic and afebrile, febrile alone, and febrile with other symptoms. We compared symptoms and viral load in infections caused by Delta and Omicron BA.1, BA.2 and BA.5 following two- or three-dose vaccination. We locked the dataset for this analysis on 28th September 2022 and censored any infection episodes 14 days prior, to mitigate against recent missing symptom diaries.

**Figure 1:**
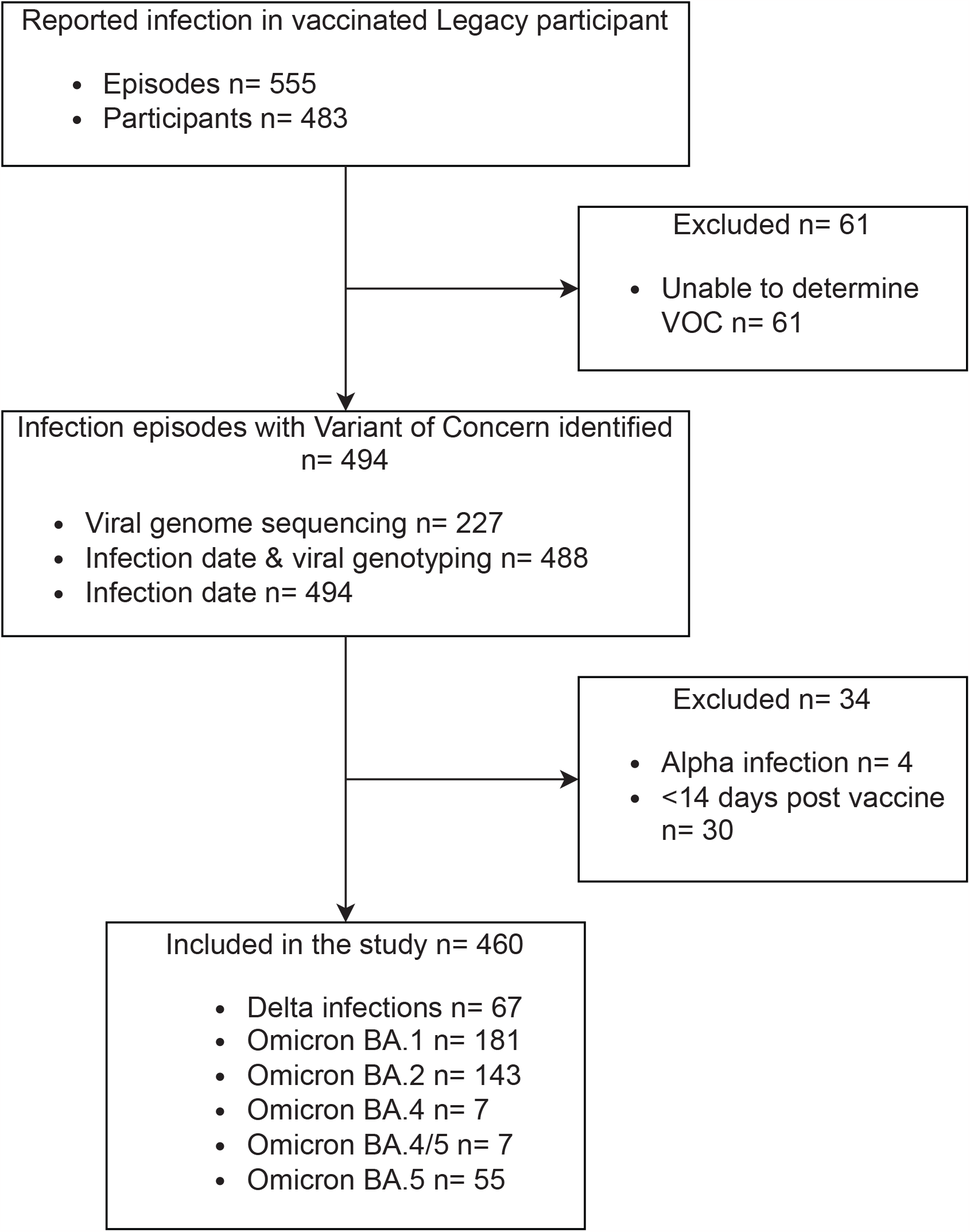
CONSORT Diagram.

### SARS-CoV-2 RT-qPCR and sequencing

RNA was extracted from self-performed upper respiratory tract swabs taken at time of breakthrough infection, as previously described (22). Viral RNA was genotyped by RT-qPCR (TaqPath COVID-19 CE-IVD Kit, ThermoFisher) to confirm SARS-CoV-2 infection. Viral RNA from positive swabs was prepared for whole-genome sequencing using the ARTIC method (https://www.protocols.io/view/ncov-2019-sequencing-protocol-v3-locost-bh42j8ye) and sequenced on the ONT GridION platform to >30k reads / sample. All swab processing for all study sites was performed in the same laboratory. The data were demultiplexed and processed using the viralrecon pipeline (https://github.com/nf-core/viralrecon).

### Data analysis, statistics, and availability

Study data were collected and managed using REDCap electronic data capture tools hosted at University College London. Data are exported weekly from REDCap into R for rollling linkage with laboratory data, visualisation and analysis(25). For this study, data were exported up to 28th September 2022 and the subsequent R record was locked. Summary descriptions of the clinical cohort and of reported symptoms and measured viral loads were generated, specifying calculation of median and IQR for continuous variables. We excluded infection episodes from analyses if their infection was ≤ 14 days after a vaccination, or if < 14 days had passed between the infection date and date of data export. For each infection episode, whole genome sequencing or a combination of date of infection and viral genotype was used to assign the VOC that caused the infection. Infection episodes were grouped by VOC and participant’s number of vaccinations and days since last vaccine dose. For the duration of symptoms and time-since-dose comparisons, infection episodes were grouped as above, and an unpaired two-tailed Wilcoxon test performed.

SARS-CoV-2 PCR data were analysed using the Cycle Threshold (Ct) of the ORF1ab gene target; smoothed spline fits were applied to Ct trajectories of all participants for each VOC. A correction of -1d was applied to original surveillance tests (but not serial swabs), assuming most surveillance tests were taken on the preceding evening. Peak Cts were drawn from the lowest Ct value (corresponding to the highest viral load) obtained from each participant between days 1-4. Ct values were compared using an unpaired two tailed Wilcoxon test.

Graphs were generated using the *ggplot2* package in R. All data (anonymised) and full R code to produce figures and statistical analysis presented in this manuscript are freely available online on Github: https://github.com/davidlvb/Crick-UCLH-Legacy-Symptoms-2022-03

### Ethical approvals

The *Legacy* study was approved by London Camden and Kings Cross Health Research Authority Research and Ethics committee (IRAS number 286469) and is sponsored by University College London Hospitals.

## Results

Infection following vaccination was reported in 555 episodes across 483 participants, resulting in a total of 1067 swabs analysed, with a median of 4 swabs per participant per infection episode. We were able to confidently determine the VOC that caused the infection in 494/555 (92%) of episodes using a combination of methods: by infection date relative to the dominant circulating VOC (494/555, 89%), a combination of infection date and viral genotyping (488/555, 8 8%), confirmed by viral genome sequencing (227/555, 41%) (**Figure 1**). We were unable to resolve the VOC in 61/555 episodes (11%) due to inconsistencies in genomic data and overlapping periods of VOC dominance; these episodes were excluded from analysis. Cases within 14 days of vaccination did not meet the definition of post-vaccine infection and were excluded (30/555, 5%), as were four alpha infections (two determined by date; two by SGTF and date). The remaining 460 episodes, across 433 individuals, were then analysed **(Table 1)**. These individuals had the same age distribution (median 39 years [IQR 31-49], as the whole *Legacy* study (40 years [31-50]), and were gender matched to *Legacy* (68 vs 68% female). Symptom questionnaires were completed for 432/460 episodes (94%). We then stratified cases into five cohorts according to the dominant combinations of participant vaccination status and virus variant (**Figure 2A**): Delta infection following 2 doses (2d+Delta: n=60, occurring a median of 155 [110-192] days since last vaccine), Delta following 3 doses (3d+Delta: n=7, 41 [26-56] days), Omicron BA.1 following 2 doses (2d+BA.1:n=27, 207[166-263] days), Omicron BA.1 following 3 doses (3d+BA.1:n=154, 82 [52-106] days, Omicron BA.2 following 3 doses (3d+BA.2: n=142, 145 [103-177] 2d+BA.2: n=1), Omicron BA.5 following 3 doses (3d+BA.5: n=55, 240 [202-276] days). Small numbers of either proven Omicron BA.4 (n=7) or indeterminate BA.4/5 infections (n=7) were also recorded, ranging between 188-474 days post last dose.

**Figure 2:**
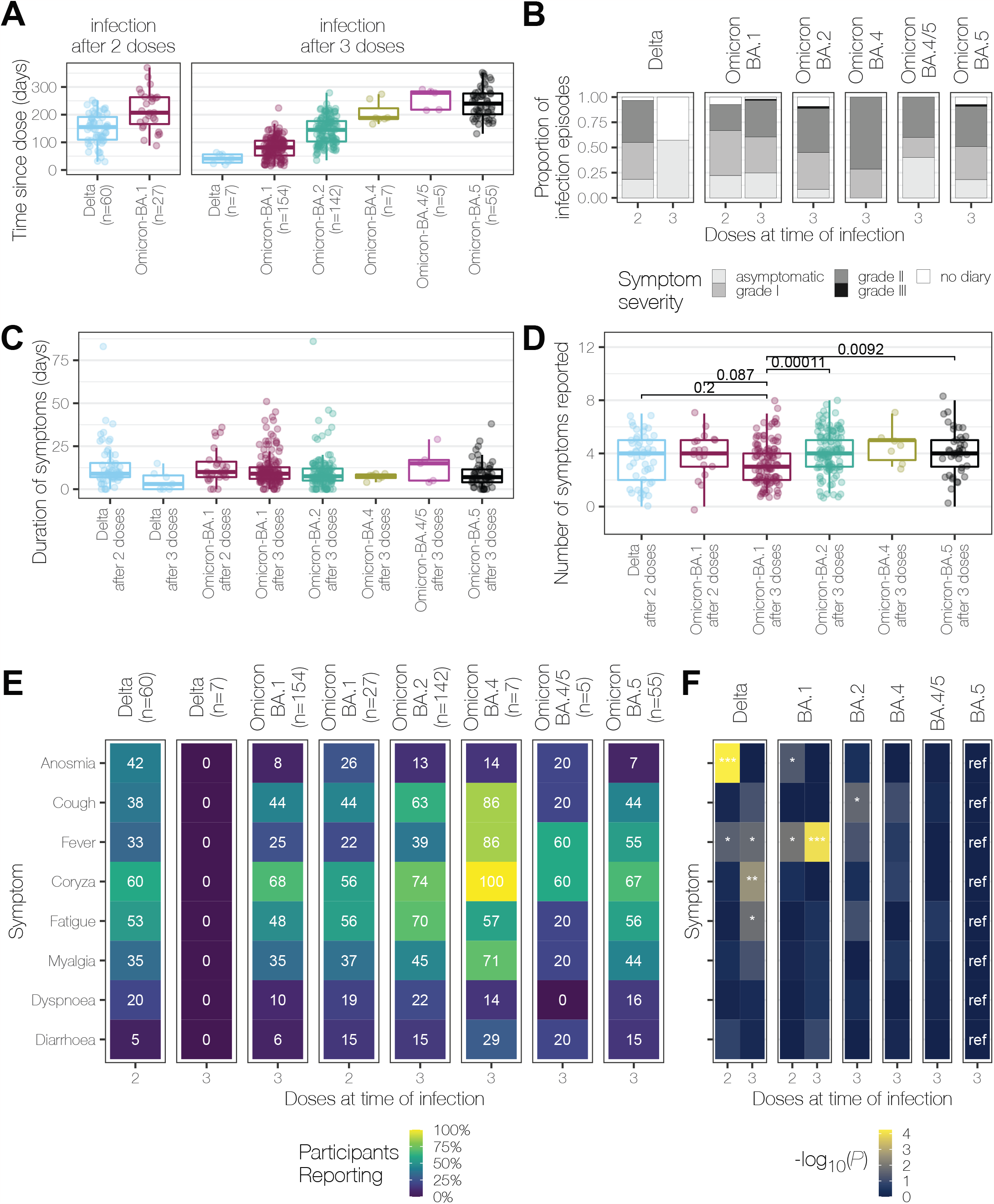
Symptoms of COVID-19 are an interaction between prevailing variants and vaccinations. **(A)** Time in days after vaccine dose before the start of an infection episode is shown for infections 14d or more after doses 2 or 3 by each VOC. **(B)** Proportion of participants reporting no, mild, moderate, or severe symptoms during their infection episode. **(C)** Duration of each infection episode in days stratified by the VOC and number of doses received prior to infection. **(D)** Number of symptoms reported by participants, stratified by VOC and number of doses received prior to infection **(E)** Percentage of individuals reporting each symptom is shown as a heatmap. Percentage shown in each tile, with the tiles shaded to reflect that percentage. The denominator used is all infection episodes of the corresponding VOC and number of doses **(F)** Heatmap showing negative decimal logarithms of P values from χ^2^ tests comparing the presence/absence of a symptom between 3d-BA.5 (ref, reference) and the indicated infection episodes. Symptoms are ordered as in Figure 2E. Significant comparisons are marked as follows: P < 0.001 with ***; P < 0.01 with ** and P<0.05 with *

While asymptomatic infections were reported in each cohort, ranging from 4/4 (100%) in 3d+Delta to 12/129 (9%) in 3d+BA.2 Omicron, the majority of participants (343/460, 75%) reported grade I-II severity illness. We found the proportion of BA.1 compared to BA.2 participants with asymptomatic infection was significantly lower (BA.1 25% asymptomatic [44/176], BA.2 9% asymptomatic [12/129]; χ ^2^ test p=0.0003 (**Figure 2B**). BA.5 was similar to BA.1 with 18% of participants reporting asymptomatic infection [10/55], χ^2^ test vs BA.1 p=0.44; vs BA.2 p= 0.09.

Within the symptomatic participants, we compared both the duration and symptom number between the cohorts. In the symptomatic cohort, the median duration of symptoms was 8 [IQR 6-13] days (**Figure 2C**). Symptom duration did not differ among any of the groups. The median number of symptoms experienced, 4(1-9) was also similar (**Figure 2D**).

However, there were significant changes in the reporting of anosmia between groups. Anosmia was reported in significantly fewer cases in the d3+Omicron cohorts (7-13%) as compared to d2+Delta (42%, P= 1.31e-08 or 1.03e-05 or 5.63e-05; χ^2^ test d2+Delta vs. Omicron BA.1 or vs. BA.2, or BA.5, respectively) (**Figure 2E**). In addition to this anosmia was less often seen in those with 3 rather than 2 doses of vaccine when infected by Omicron BA.1 but not Delta (26% vs 8%, p = 0.01 or 40% vs 0% p = 0.08; χ ^2^ test for dose 2 or 3 at time of infection with Omicron BA.1 or Delta respectively).

While cough was reported slightly more frequently in d3+Omicron BA.1 as compared to d2+Delta, and slightly less frequently in d3+Omicron BA.5, neither difference was significant (44% vs. 38%, p=0.59, or 44% vs. 38%, p=0.70 respectively; χ ^2^ tests). Cough was more frequently reported in 3d+BA.2 compared to d2+Delta (63% vs. 38%, P=0.02 χ ^2^ test). Indeed, despite reports of changes in tissue tropism in laboratory studies of Omicron BA.1(26),the proportion of participants reporting coryza, fatigue, myalgia, fever, shortness of breath, and diarrhoea remained broadly similar across all combinations of cohorts that reported symptomatic illness (d2+Delta, d2+BA.1, d3+BA.1, BA.2, BA.4 and BA.5). Fever was a common feature of 3d+BA.5 infections, with 55% of symptomatic infections including self-reported fever, compared to 33% for 2d+Delta (p=0.04), 25% for 3d+BA.1 (p=0.09×10^−3^ and 39% for 3d+BA.2 (p=0.06, χ ^2^ tests).

We undertook a hierarchical clustering analysis of the symptom data to investigate which symptoms presented simultaneously **(Figure 3)**. Overall, symptom clusters did not associate with variants (**Figure 3A**). However, when symptoms were analysed for individual VOCs, we found some distinct patterns. While both coryza and fatigue clustered together in Delta infections, cough and fever were less likely to be reported together; myalgia was reported in a minority of cases (around one-third), and clustered with fever (**Figure 3B)**. Symptoms caused by infections with Omicron BA.1 and BA.2 were dominated by clusters consisting of cough, coryza and fatigue; fever and myalgia were less common but did predominately occur in this same cluster **(Figures 3C,E)**. In contrast, participants with BA.5, 4/5 and BA.4 infections most frequently reported coryza, with fever, myalgia, and fatigue most commonly co-reported (**Figures 3D,F,G**). Of patients self-reporting fever, almost all experienced another symptom, with only 2 individuals reporting fever alone (one after d3+BA.1, and one after d2+Delta).

**Figure 3:**
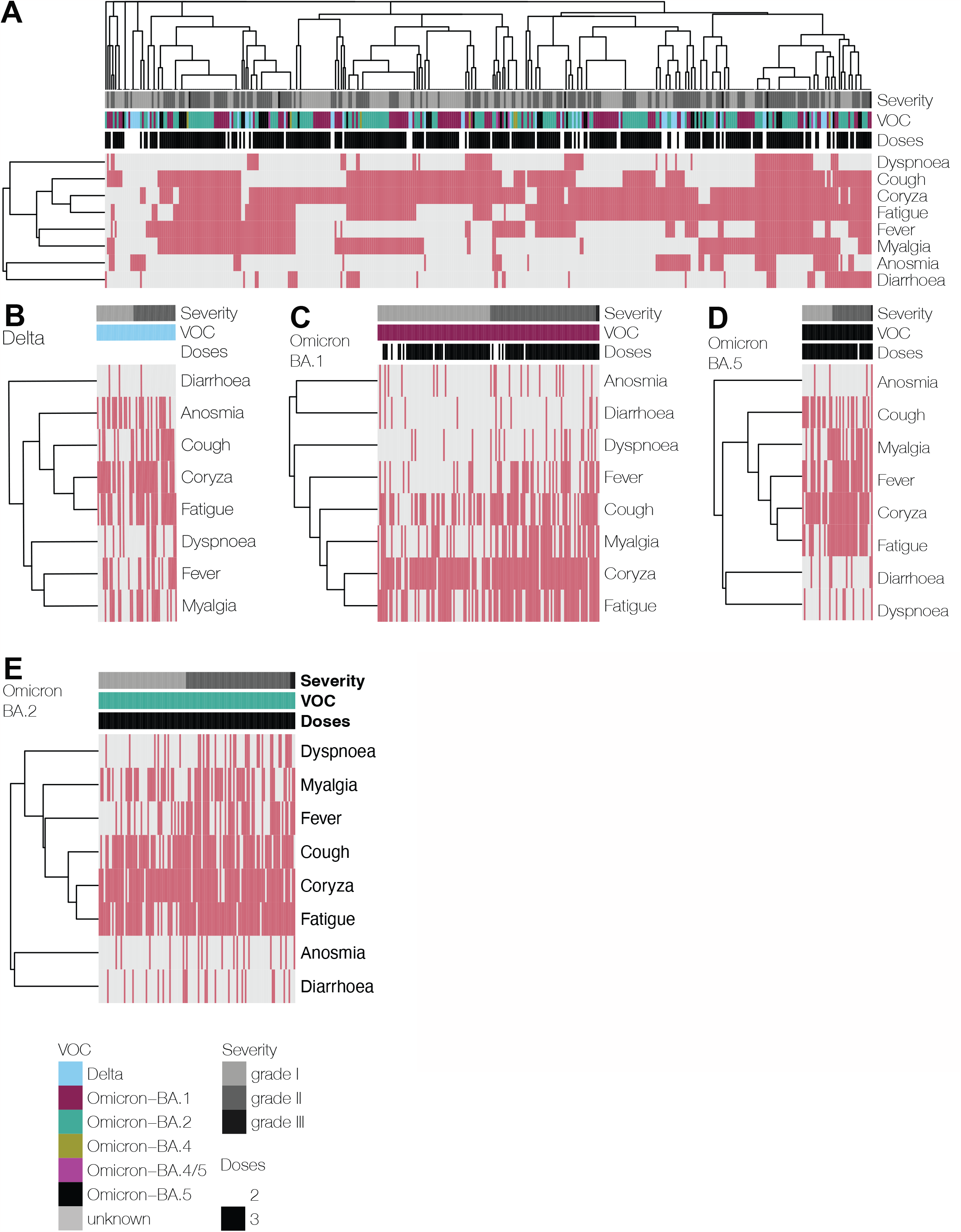
Hierarchical clustering of symptom patterns in infection episodes occurring after dose 2. All symptomatic episodes from the Legacy cohort clustered by individual (columns) and symptoms (rows) using Jaccard distances. The presence of a symptom is indicated by rose shading and the absence of a symptom by grey shading. Above each individual episode (each column) the colour bar indicates the severity, assigned VOC of that infection episode, and the number of doses of vaccine received before that infection. An individual may be present >1 if they experienced more than one infection episodes. Symptomatic episodes depicted across all VOCs in **(A)**, all infection episodes, **(B)** Delta, **(C)** Omicron BA.1 **(D)** Omicron BA.5, **(E)** Omicron BA.2. Asymptomatic infection episodes are not shown.

While most participants were not febrile (75% or 61% for d3+BA.1 or d3+BA.2), NHS guidance also recommends self-isolation if an individual feels too unwell to carry out their routine activities. On the assumption that those reporting moderate severity symptoms (grades II or III) would not be able to attend work under the April 2022 guidelines(24) and therefore would self-isolate, 44% of our cohort with active infection (3d+BA.1 [38%] or 3d+BA.2 [50%] 3d+BA.5 [45%]) would still not meet self-isolation criteria for either for fever or severity, and thus would enter social circulation whilst likely infectious.

To test if symptoms were associated with viral replication, we examined infection dynamics in more detail. Participants who reported acute infection provided serial self-performed upper respiratory tract swabs for RT-qPCR analysis of SARS-CoV-2 RNA during isolation, with predominately Delta, BA.1 and BA.2 infections. We profiled the kinetics of each infection using the Ct value, as an inverse proxy for representative of levels of replicating, viable virus (27, 28). Across all cohorts, the median Ct values remained at levels considered to be infectious for 7-10 days, irrespective of symptom severity and including asymptomatic participants (**Figure 4A**). The lowest Ct values (corresponding to estimated peak viral load (27), hereafter referred to as peak) were observed between 2-5 days after symptom onset, with similar Ct trajectories observed across all VOCs tested. The only observed significant difference in the peak Ct value was between those who experienced BA.2 infection (median minimum Ct 18.0) following 3 doses as compared to BA.1 (median minimum Ct 20.9, unpaired two tailed Wilcoxon test, p=0.0016) **(Figure 4B)**. Peak Ct values did not differ between other VOC tested including Delta and Omicron BA.4/5. We then examined if dynamic and peak Ct values differed between those with fever and those who were symptomatic though afebrile. We found virtually identical viral load trajectories in those participants where we had adequate serial sampling Omicron BA.1 and BA.2 (**Figure 4C**).

**Figure 4:**
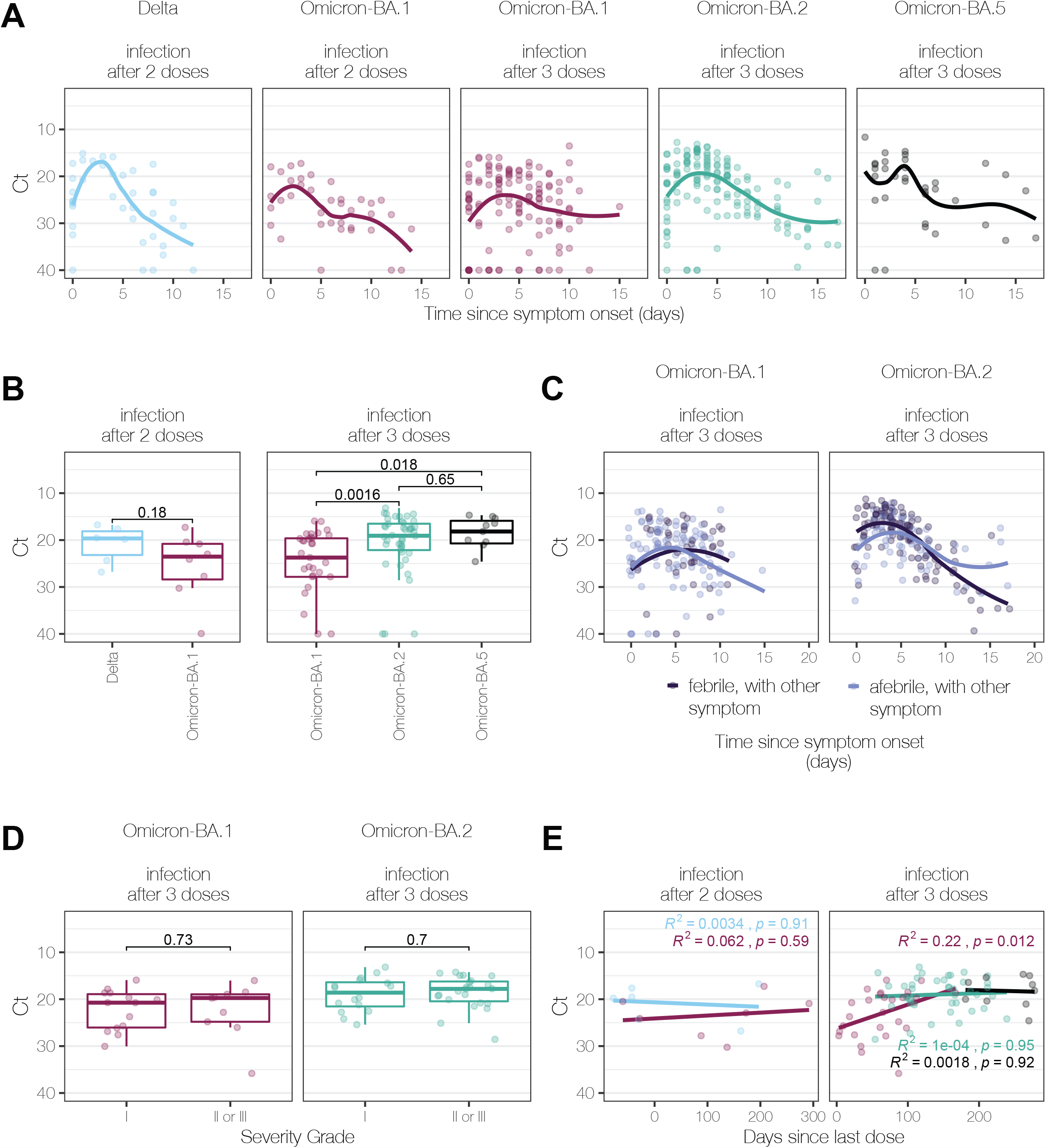
Peak viral load from symptomatic infection episodes in triple-vaccinated participants, compared to days since vaccination. **(A)** Viral load (Ct) trajectories (day 0 = symptom onset), plotted separately for each variant and stratified by the number of preceding vaccinations. Smoothed spline fits are shown. **(B)** Peak viral load on days 1-4 following symptom onset from Delta, Omicron BA.1, BA.2 and BA.5 infection episodes by vaccine dose number. **(C)** Viral load (Ct) trajectories for symptomatic BA.1 or BA.2 infections with febrile and afebrile infection episodes in dark or light blue respectively. Smoothed spline fits are shown. **(D)** Peak viral load on days 1-4 in participants with either BA.1 or BA.2 reported by symptom severity grade. **(E)** Peak Ct value across Delta, BA.1, BA.2 and BA.5 plotted against the time in days since last vaccine dose after either two or three vaccine doses. Lines from linear regression are shown, with Spearman’s R_2_ and P values shown.

Notably, there was also no significant difference in lowest Ct values in those who met the updated NHS criteria for isolation and those who did not. Amongst symptomatic individuals, the presence of fever did not significantly affect the lowest Ct value for either BA.1 or BA.2 infections: median minimum Ct febrile vs. afebrile BA.1 (21.1 vs 22.0, p=0.28) and BA.2 (17.9 vs 19.8, p=0.16) **(Figure 4C)**. Furthermore, the self-reported severity grade was not associated with differences in lowest Ct measurements for either BA.1 (lowest Ct grade I vs grades II-III = 20.7 vs 24.8, p=0.73) or BA.2 (lowest Cts grade I vs grades II-III = 20.1 vs 18.0, p=0.7) **(Figure 4D)**. We did detect a trend towards lower Ct and increasing time since last vaccination dose that was significant for BA.1 infection after 3 doses (R^2^ = 0.22 p=0.012), but this did not reach statistical significance for BA.2 (R^2^ =0.0001 p=0.95) or BA.5 (R^2^=0.0018 p=0.92) (**Figure 4E**).

## Discussion

Our large, longitudinal cohort study demonstrates the evolution of symptom profiles between Delta and Omicron sub-variants, including Omicron BA.5. In contrast, we also show the relatively unchanging peak viral loads in the respiratory tract, regardless of variant or vaccine history, with implications for ongoing SARS-CoV-2 transmission. Despite analyses of hospitalisation and mortality data indicating that Omicron caused less-severe clinical disease (8, 9, 14), our data suggests that COVID-19 caused by all Omicron sub-variants continues to cause a significant symptom burden in non-hospitalised adults, with attendant impacts on healthcare resources and the economic impact of increased time off due to illness. Continued infections with new variants, despite a vaccinated population, are therefore likely to continue to contribute to increasing incidence of post-COVID syndrome (PCS) or Long COVID (16-18).

Understanding the symptomatology of SARS-CoV-2 variants is critical, in order to direct public health messaging and testing guidance. We show anosmia, a key and relatively specific symptom of earlier SARS-COV-2 variants, is less common across the Omicron sub-variants to BA.5 than in Delta. This replicates the findings of large community based studies in the UK (ZOE, REACT) and the USA (19, 21, 29, 30), where reduction in anosmia incidence is the most notable difference between Delta and Omicron infections to BA.2. Anosmia caused by SARS-CoV-2 is thought to be due to downregulation or damage to receptors in olfactory epithelial supporting sustentacular cells (31). Omicron is postulated to have reduced tissue tropism to these cells in the nasal epithelium to Delta, due to differential utilisation of transmembrane serine protease 2 (TMPRSS2)(32). TMPRSS2 is a key host protein for virus entry in Delta, mechanistic reports suggest viral damage to sustentacular cells by Delta and earlier VOC is the mechanism mediating anosmia, subsequently attenuated by Omicron sub-variants that do not utilise this pathway for replication to the same degree (33, 34). We also found that anosmia was less likely to be reported by participants after three doses of vaccines rather than two during Omicron sub-variant infections, other studies do not report incidence of anosmia in relation to vaccine doses. We hypothesise that accumulating vaccine doses may temporarily increase mucosal IgA and neutralising antibodies, further attenuating the damaging effects of viral replication in the olfactory cells (35, 36). However, other upper respiratory symptoms, other than anosmia, and associated viral kinetics remained unchanged across variants irrespective of vaccine status, and thus it is unlikely that vaccine-induced mucosal antibodies significantly affect viral replication across the wider nasal epithelial surface.

Fever has been shown in other studies to be less frequently reported in infection episodes with Omicron BA.1/2 (19, 30). We replicated this finding in our study, however, we found fever frequency increased in Omicron BA.5 infections when compared to Delta infections. Other studies reporting variant-specific symptom profiles have not extended reporting to BA.5 infections, including the REACT study, which reported reduced fever incidence in BA.1 and BA.2 (30); and the app-based ZOE study, with participants reporting decreased fever associated with Omicron BA.1 infections only compared to Delta (19). Our finding of increased fever with BA.5 infections is in line with prospective population level data recently reported from Japan(37). Why fever may be more common with BA.5 infections is unknown, but waning immunity and increasing antigenic divergence may both impact on increasing symptom severity.

Similarly to these large, population based studies, we found a non-significant trend towards increasing coryza symptoms in both BA.2 and BA.5 infections. However these larger studies were able to analyse specific upper respiratory symptoms, where participants reported higher rates of symptoms in keeping with coryza in Omicron BA.1 and BA.2, with ‘sneezing’, ‘sore throat’ and ‘runny nose’ reaching significance individually(30) and the ZOE study where ‘sneezing’ and ‘runny nose’ were associated with Delta infections, whereas ‘sore throat’ and ‘hoarse voice’ were more likely to occur in Omicron BA.1 infections (19, 30). Reporting of coryza, or ‘cold like symptoms’ alone is potentially subject to different interpretation by participants, and thus too broad a category to distinguish between variants and sub-variants in relatively smaller studies such as ours.

Due to the nature of occupational health PCR screening in the *Legacy* cohort, we were able to include true asymptomatic infections within our analysis, contrasting with cohort studies relying on symptom-triggered testing(19, 30, 37). We found all cases of Delta infection following three vaccine doses were asymptomatic, contrasting with participants infected with Omicron sub-variants, who were more likely to be symptomatic despite a similar 3-month interval since vaccination and near-identical viral load trajectories across VOCs. The Delta comparator group was relatively small, but our data suggest that the immediate boosting effect of third vaccine dose may minimise symptoms, but this protection is short-lived. We found more individuals were vulnerable to symptomatic disease at the longer post-dose intervals when BA.2 and then BA.4/5 emerged which may be related to waning of vaccine-induced immunity (38, 39),(19). Furthermore, we show an association between increasing time since vaccination and increasing viral loads in BA.1 infections, not captured by previous studies, that suggests waning mucosal, as well as humoral immunity may be exploited by SARS-CoV-2 (19, 20, 35).

Using data from both prior negative screening tests, and symptomatic onset, we were able to show peak Ct, and therefore presumed peak infectivity, occurred 2-5 days after symptoms onset, in line with other work capturing known viral trajectory studies (27, 40) and indicative of ongoing infectivity up to 7-10 days across all variants tested (41-43).

It is not fully clear from our data to what extent vaccination may suppress viral replication and transmission. We observed almost identical viral load trajectories across VOCs, irrespective of vaccination status and time since vaccination, which corresponded closely with those found during both controlled human challenge models in unvaccinated individuals, asymptomatic household transmission in South Africa and healthcare workers in Turkey (20, 27, 28), suggesting that immunity induced by first-generation vaccines encoding an ancestral Spike might have minimal impact on viral replication in the nasopharynx. However, we also observed a trend towards higher peak viral loads after longer time since vaccination that reached significance in BA.1 infections, mirroring waning neutralising antibodies in vaccinated cohorts (44). Coupled with the similarity in symptom profiles between those infected with Omicron sub-variants BA.1, BA.2 and BA.4/5, waning of neutralising antibodies induced by vaccination may be marginally implicated in viral clearance, in parallel with increasing risk of hospitalisation (44-47). Such results would also be consistent with reduced transmission inferred from household attack rates in vaccinated compared to unvaccinated cohorts (48).

The reported shift in COVID-19 symptoms between Delta and Omicron sub-variants may be driven by intrinsic differences in the biology of the virus itself, leading to modified host responses; mutations in the Spike protein of SARS-CoV-2 VOCs may directly impact on symptom profiles through changes in viral tropism (2-4, 49, 50). Detailed data on differences in the host response to different VOC in vaccinated, non-hospitalised adults are currently lacking, but are likely to involve an interplay between localised mucosal innate and early T-cell interferon-mediated systemic responses (2, 51, 52). Further studies to determine the mechanisms by which different and emerging VOCs interact with the mucosal and systemic response, are urgently required and will be a necessary counterpart to clinical studies in the assessment of future VOCs.

In April 2022 the UK Government updated their self-isolation and testing advice to recommend a symptom-based approach aiming to identify people more likely to be infectious. However, we found the majority of participants infected with Omicron sub-variants were afebrile, and almost half were asymptomatic or had mild symptoms, and therefore did not meet this guidance for either testing or isolation. Combined with the removal of free testing, UK population estimates of COVID-19 across the Omicron BA.2 and BA.5 waves in 2022 are therefore likely to be subject to significant under-ascertainment. While our study does not examine transmission directly, it is notable that nearly all participants had low Ct values corresponding with high viral loads in the nasopharynx that tended to remain <25 beyond the isolation period recommended by many countries, including the UK and US (1, 24). Many participants in our study were thus potentially infectious beyond current isolation periods and thus present an ongoing risk to CEV individuals particularly. Our study, with others suggests that as Omicron continues to evolve, and continues to cause a significant burden of disease for clinically vulnerable adults, isolation and testing guidance should remain under regular review (53-55). We suggest that updated isolation guidance for each emerging VOC needs to be informed by data on symptoms, antibody trajectories and viral kinetic data, incorporated into mathematical models of transmission and population behaviour (56).

Such updated isolation guidance would also need to incorporate factors beyond those addressed in this study, notably data from younger & older age groups, from the substantial, understudied minority who remain unvaccinated (6.1% of the English population) and those eligible for the bivalent mRNA vaccine (13). Due to the rapid evolution of SARS-CoV-2 across 2022, in combination with changing national vaccination policy, our data are subject to important limitations. While we were able to obtain detailed prospective clinical and PCR data from our cohort across four waves of infection, we were not able to compare these data with earlier waves (i.e. Alpha/B.1.1.7, EU1/B.1.177, and D614G/B.1) as these preceded our study period of enhanced infection surveillance. We were unable to control for differences in clinical baseline demographics between infection groups, however *Legacy* participants are representative of healthy working age adults in London, less than 10% of our cohort have significant clinical co-morbidities (5).

The viral load trajectories we observed were remarkably consistent and closely mirrored supervised testing in the SARS-CoV-2 human challenge study (27), suggesting high quality samples were obtained at serial time points across participants, however serial PCR testing was not supervised by a clinician. Participants were given clear instructions on testing and had been compliant with asymptomatic screening for excess of 12 months prior to the start of this study, sampling variability is likely to have a minimal effect on our results (57). Similarly, while symptom profiles and severity were self-reported during infection, all diaries were checked for accuracy with the participant by a study clinician within 21 days of reported symptom onset to minimise recall bias.

The Ct assay used was validated in house against known viral copy numbers, obtained from live-virus quantification (5, 22). Our reported Ct values are consistent with Omicron in the general population (58), and are well within the range in which infectious virus could be detected during both human challenge with SARS-CoV-2 and other prospectively sampled cohorts (27, 28, 59). The heterogenity in lowest value and longditinal Ct kinetics in our cohort are also very similar to other cohorts (42, 60).

In conclusion, we show that symptoms experienced by vaccinated adults are likely to change with new SARS-CoV-2 VOCs including within defined lineages such as Omicron BA.1-5. Guidance on self-isolation and testing requires regular evaluation from prospective clinical studies as new VOCs emerge, and notably, neither symptom severity nor presence of fever is a useful proxy for testing when considering the need for self-isolation. Updated advice should continue to emphasise the ongoing risk of transmission from individuals with no or mild symptoms whilst infected with Omicron to vulnerable populations, and the possibility for characteristic symptoms to change in the future were a new VOC to emerge.

## Data Availability

All data produced in the present work are contained in the manuscript and will be made openly available online after peer review

## Role of the funding source

This work was supported by UCLH/UCL who received a proportion of funding from the National Institute for Health Research (NIHR) University College London Hospitals Department of Health’s NIHR Biomedical Research Centre (BRC). EW, VL and BW are supported by the Centre’s funding scheme. This work was supported jointly by the BRC and core funding from the Francis Crick Institute, which receives its funding from Cancer Research UK, the UK Medical Research Council, and the Wellcome Trust. This research was funded in whole, or in part, by the Wellcome Trust [FC011104, FC011233, FC001030, FC001159, FC001827, FC001078, FC001099, FC001169, 206250/Z/17/Z]. For the purpose of Open Access, the author has applied a CC BY public copyright licence to any Author Accepted Manuscript version arising from this submission. DLVB is additionally supported by the Genotype-to-Phenotype National Virology Consortium (G2P-UK) via UK Research and Innovation and the UK Medical Research Council, and TWR and AJK are additionally supported by the Wellcome Trust. The funders of the study had no role in study design, data collection, data analysis, data interpretation, or writing of the report. The corresponding authors had full access to all the data and the final responsibility to submit for publication.

## Declaration of interests

CSw reports interests unrelated to this Correspondence: grants from BMS, Ono-Pharmaceuticals, Boehringer-Ingelheim, Roche-Ventana, Pfizer and Archer Dx, unrelated to this Correspondence; personal fees from Genentech, Sarah Canon Research Institute, Medicxi, Bicycle Therapeutics, GRAIL, Amgen, AstraZeneca, BMS, Illumina, GlaxoSmithKline, MSD, and Roche-Ventana, unrelated to this Correspondence; and stock options from Apogen Biotech, Epic Biosciences, GRAIL, and Achilles Therapeutics, unrelated to this Correspondence. DLVB reports grants from AstraZeneca unrelated to this Correspondence. All other authors declare no competing interests.

## Acknowledgements

The authors would like to thank all the study participants, the staff of the NIHR Clinical Research Facility at UCLH including Kirsty Adams and Marivic Ricamara. We would like to thank Jules Marczak, Gita Mistry, and the staff of the Scientific Technology Platforms (STPs) and COVID-19 testing pipeline at the Francis Crick Institute.

## Notes

### Author Declarations

The Legacy study was approved by London Camden and Kings Cross Health Research Authority Research and Ethics committee (IRAS number 286469)

### Summary of Updates

corrected space in the title

