## Supplemental Table 1 for "COVID-19 in non-hospitalised adults caused by either SARS-CoV-2 sub-variants Omicron BA.1, BA.2, BA.5 or Delta associates with similar illness duration, symptom severity and viral kinetics, irrespective of vaccination history"

| **Characteristic** | **Delta (2)**  N = 60 | **Delta (3)**  N = 7 | **Omicron-BA.1 (2)**  N = 27 | **Omicron-BA.1 (3)**  N = 154 | **Omicron-BA.2 (2)**  N = 1 | **Omicron-BA.2 (3)**  N = 142 | **Omicron-BA.4 (3)**  N = 7 | **Omicron-BA.4/5 (2)**  N = 2 | **Omicron-BA.4/5 (3)**  N = 5 | **Omicron-BA.5 (3)**  N = 55 |
| --- | --- | --- | --- | --- | --- | --- | --- | --- | --- | --- |
| Dose 2 |  |  |  |  |  |  |  |  |  |  |
| AZD1222 | 30 (50%) | 0 (0%) | 11 (41%) | 35 (23%) | 1 (100%) | 35 (25%) | 3 (43%) | 0 (0%) | 0 (0%) | 17 (31%) |
| BNT162b2 | 29 (48%) | 7 (100%) | 13 (48%) | 115 (75%) | 0 (0%) | 99 (70%) | 4 (57%) | 2 (100%) | 5 (100%) | 36 (65%) |
| mRNA1273 | 1 (1.7%) | 0 (0%) | 3 (11%) | 4 (2.6%) | 0 (0%) | 7 (4.9%) | 0 (0%) | 0 (0%) | 0 (0%) | 2 (3.6%) |
| Other | 0 (0%) | 0 (0%) | 0 (0%) | 0 (0%) | 0 (0%) | 1 (0.7%) | 0 (0%) | 0 (0%) | 0 (0%) | 0 (0%) |
| Dose 3 |  |  |  |  |  |  |  |  |  |  |
| AZD1222 | 0 (0%) | 0 (0%) | 0 (0%) | 1 (0.6%) | 0 (NA%) | 0 (0%) | 0 (0%) | 0 (NA%) | 0 (0%) | 0 (0%) |
| BNT162b2 | 39 (76%) | 7 (100%) | 12 (92%) | 142 (92%) | 0 (NA%) | 136 (96%) | 5 (71%) | 0 (NA%) | 5 (100%) | 48 (87%) |
| mRNA1273 | 12 (24%) | 0 (0%) | 1 (7.7%) | 10 (6.5%) | 0 (NA%) | 6 (4.2%) | 2 (29%) | 0 (NA%) | 0 (0%) | 7 (13%) |
| Others | 0 (0%) | 0 (0%) | 0 (0%) | 1 (0.6%) | 0 (NA%) | 0 (0%) | 0 (0%) | 0 (NA%) | 0 (0%) | 0 (0%) |
| Unknown | 9 | 0 | 14 | 0 | 1 | 0 | 0 | 2 | 0 | 0 |
| Site |  |  |  |  |  |  |  |  |  |  |
| CNWL | 0 (0%) | 1 (14%) | 0 (0%) | 3 (1.9%) | 0 (0%) | 4 (2.8%) | 0 (0%) | 0 (0%) | 0 (0%) | 3 (5.5%) |
| Crick (non-NHS) | 46 (77%) | 4 (57%) | 24 (89%) | 99 (64%) | 1 (100%) | 107 (75%) | 6 (86%) | 0 (0%) | 2 (40%) | 42 (76%) |
| Ealing/NWP | 2 (3.3%) | 1 (14%) | 1 (3.7%) | 2 (1.3%) | 0 (0%) | 5 (3.5%) | 0 (0%) | 1 (50%) | 0 (0%) | 0 (0%) |
| UCLH | 12 (20%) | 1 (14%) | 2 (7.4%) | 50 (32%) | 0 (0%) | 26 (18%) | 1 (14%) | 1 (50%) | 3 (60%) | 10 (18%) |
| Sex |  |  |  |  |  |  |  |  |  |  |
| Female | 37 (62%) | 4 (57%) | 17 (63%) | 107 (69%) | 0 (0%) | 97 (68%) | 6 (86%) | 2 (100%) | 5 (100%) | 40 (73%) |
| Male | 23 (38%) | 3 (43%) | 10 (37%) | 47 (31%) | 1 (100%) | 45 (32%) | 1 (14%) | 0 (0%) | 0 (0%) | 15 (27%) |
| Median age (years) [IQR] | 38 [28-48] | 47 [40-52] | 34 [29-40] | 40 [32-49] | 36 [36-36] | 40 [31-48] | 32 [26-40] | 52 [52-52] | 57 [44-57] | 37 [29-50] |
| Episode number |  |  |  |  |  |  |  |  |  |  |
| 1 | 60 (100%) | 6 (86%) | 25 (93%) | 149 (97%) | 1 (100%) | 119 (84%) | 6 (86%) | 1 (50%) | 3 (60%) | 33 (60%) |
| 2 | 0 (0%) | 1 (14%) | 2 (7.4%) | 5 (3.2%) | 0 (0%) | 22 (15%) | 1 (14%) | 1 (50%) | 2 (40%) | 22 (40%) |
| 3 | 0 (0%) | 0 (0%) | 0 (0%) | 0 (0%) | 0 (0%) | 1 (0.7%) | 0 (0%) | 0 (0%) | 0 (0%) | 0 (0%) |
| Joined study before infection episode? |  |  |  |  |  |  |  |  |  |  |
| No | 25 (42%) | 5 (71%) | 16 (59%) | 55 (36%) | 0 (0%) | 42 (30%) | 0 (0%) | 1 (50%) | 2 (40%) | 11 (20%) |
| Yes | 35 (58%) | 2 (29%) | 11 (41%) | 99 (64%) | 1 (100%) | 100 (70%) | 7 (100%) | 1 (50%) | 3 (60%) | 44 (80%) |
| Median days since dose prior to infection [IQR] | 155 [110-192] | 41 [26-56] | 207 [166-263] | 82 [52-106] | 333 [333-333] | 145 [103-177] | 188 [184-223] | 400 [325-474] | 279 [218-284] | 240 [202-276] |
| Self-reported symptom severity |  |  |  |  |  |  |  |  |  |  |
| Grade I | 22 (38%) | 0 (0%) | 12 (48%) | 55 (36%) | 0 (NA%) | 52 (40%) | 2 (29%) | 0 (0%) | 1 (20%) | 18 (35%) |
| Grade II | 25 (43%) | 0 (0%) | 7 (28%) | 56 (37%) | 0 (NA%) | 62 (48%) | 5 (71%) | 2 (100%) | 2 (40%) | 22 (43%) |
| Grade III | 0 (0%) | 0 (0%) | 0 (0%) | 2 (1.3%) | 0 (NA%) | 3 (2.3%) | 0 (0%) | 0 (0%) | 0 (0%) | 1 (2.0%) |
| Asymptomatic | 11 (19%) | 4 (100%) | 6 (24%) | 38 (25%) | 0 (NA%) | 12 (9.3%) | 0 (0%) | 0 (0%) | 2 (40%) | 10 (20%) |
| Unknown | 2 | 3 | 2 | 3 | 1 | 13 | 0 | 0 | 0 | 4 |
| Self-reported duration of symptoms | 9 [7-15] | 3 [0-8] | 10 [7-16] | 9 [6-13] | 6 [6-6] | 8 [5-12] | 8 [6-8] | 8 [8-8] | 15 [5-17] | 7 [4-12] |

­
